# Causes, Diagnostic Testing, and Treatments Related to Clinical Deterioration Events among High-Risk Ward Patients

**DOI:** 10.1101/2024.02.05.24301960

**Authors:** Matthew M. Churpek, Ryan Ingebritsen, Kyle A. Carey, Saieesh A Rao, Emily Murnin, Tonela Qyli, Madeline K. Oguss, Jamila Picart, Leena Penumalee, Benjamin D. Follman, Lily K Nezirova, Sean T. Tully, Charis Benjamin, Christopher Nye, Emily R. Gilbert, Nirav S. Shah, Christopher J. Winslow, Majid Afshar, Dana P. Edelson

**Author notes:** Corresponding author: Matthew M. Churpek, MD, MPH, PhD. **Disclosures and conflicts of interest:** Drs. Churpek and Edelson are named inventors on a patent (#11,410,777) for eCART and receive royalties from the University of Chicago for this intellectual property. Dr. Edelson is employed by and has an equity stake in AgileMD, which markets and distributes eCART.

## Abstract

**OBJECTIVE:** Timely intervention for clinically deteriorating ward patients requires that care teams accurately diagnose and treat their underlying medical conditions. However, the most common diagnoses leading to deterioration and the relevant therapies provided are poorly characterized. Therefore, we aimed to determine the diagnoses responsible for clinical deterioration, the relevant diagnostic tests ordered, and the treatments administered among high-risk ward patients using manual chart review.

**DESIGN:** Multicenter retrospective observational study

**SETTING:** Inpatient medical-surgical wards at four health systems from 2006-2020 **PATIENTS:** Randomly selected patients (1,000 from each health system) with clinical deterioration, defined by reaching the 95th percentile of a validated early warning score, electronic Cardiac Arrest Risk Triage (eCART), were included.

**INTERVENTIONS:** None

**MEASUREMENTS AND MAIN RESULTS:** Clinical deterioration was confirmed by a trained reviewer or marked as a false alarm if no deterioration occurred for each patient. For true deterioration events, the condition causing deterioration, relevant diagnostic tests ordered, and treatments provided were collected. Of the 4,000 included patients, 2,484 (62%) had clinical deterioration confirmed by chart review. Sepsis was the most common cause of deterioration (41%; n=1,021), followed by arrhythmia (19%; n=473), while liver failure had the highest in-hospital mortality (41%). The most common diagnostic tests ordered were complete blood counts (47% of events), followed by chest x-rays (42%), and cultures (40%), while the most common medication orders were antimicrobials (46%), followed by fluid boluses (34%), and antiarrhythmics (19%).

**CONCLUSIONS:** We found that sepsis was the most common cause of deterioration, while liver failure had the highest mortality. Complete blood counts and chest x-rays were the most common diagnostic tests ordered, and antimicrobials and fluid boluses were the most common medication interventions. These results provide important insights for clinical decision-making at the bedside, training of rapid response teams, and the development of institutional treatment pathways for clinical deterioration.

**KEY POINTS:** **Question:** What are the most common diagnoses, diagnostic test orders, and treatments for ward patients experiencing clinical deterioration?

**Findings:** In manual chart review of 2,484 encounters with deterioration across four health systems, we found that sepsis was the most common cause of clinical deterioration, followed by arrythmias, while liver failure had the highest mortality. Complete blood counts and chest x-rays were the most common diagnostic test orders, while antimicrobials and fluid boluses were the most common treatments.

**Meaning:** Our results provide new insights into clinical deterioration events, which can inform institutional treatment pathways, rapid response team training, and patient care.

## INTRODUCTION

The early identification and treatment of clinical deterioration in patients outside the intensive care unit (ICU) is associated with improved outcomes (1–4). This has led to the development of early warning scores, such as the Modified Early Warning Score (MEWS) and the electronic Cardiac Arrest Risk Triage Score (eCART), which use physiological variables (e.g., vital signs, laboratory values) to predict which patients are at the highest risk of deterioration (5–7). These scores serve as the afferent detection arm of rapid response systems designed to bring additional resources to the bedside for the diagnosis and treatment of the underlying cause of clinical deterioration (8–10). Recent evidence suggests that integration of these scoring systems into the electronic health record (EHR) may improve important outcomes, such as in-hospital mortality (1, 2).

Although the predictors of clinical deterioration episodes, such as elevated respiratory rate, have been well described (11–14), the medical conditions causing these episodes as well as their related diagnostic tests and treatments are poorly characterized. Prior epidemiological studies of clinical deterioration events have been limited to patients requiring certain interventions (e.g., ICU transfer or rapid response activation) or having specific vital sign triggers (e.g., elevated respiratory rate), measures which do not generalize to all deterioration events (15–18). Because it is not the early warning scores but the process of diagnosis and treatment that they prompt that drives clinical improvement, an understanding of the most common causes of clinical deterioration would allow for the development of more targeted workflows. For example, this knowledge could help hospitals prioritize the most common conditions or those with the highest mortality rate when considering what screening questionnaires (e.g., sepsis screening), diagnostic tests, and clinical actions should be prioritized in these workflows. It could also assist with determining what resources may be needed during clinical deterioration events.

Therefore, we aimed to determine the underlying cause of clinical deterioration for high-risk ward patients outside the ICU as well as the most commonly ordered diagnostic tests and treatments. To do this, we performed detailed manual chart review with expert annotation at four health systems and determined the medical condition causing the deterioration episode. We then compared patient characteristics and outcomes across the different conditions, including whether these conditions were present on admission and each condition’s associated in-hospital mortality. Finally, we collected the diagnostic tests and interventions related to these deterioration events.

## MATERIALS AND METHODS

### Study Design and Population

All adult (age ≥18 years) hospitalized patients admitted to the University of Chicago Medicine, the University of Wisconsin-Madison Hospital, Loyola University Medical Center, and four NorthShore University HealthSystem hospitals spanning 2007-2020 were eligible for inclusion in this observational retrospective study. Patients were excluded if no clinician provider notes (e.g., Admission History and Physical, Discharge Summary, etc.) were available during the admission or if they were never admitted to the medical-surgical (non-ICU) wards. eCART scores, which are the probability of a patient experiencing a cardiac arrest, ICU transfer, or death within the next eight hours, were calculated for all patients during their ward stay using demographics, vital signs, and laboratory results using a previously published model (5). A randomly selected 1,000 encounters from each of the four health systems that reached the 95^th^ percentile of the eCART score, with the threshold determined using data from all ward observations at the University of Chicago where the chart reviews were first started, were included in the study for manual chart review (4,000 admissions total). The study was approved by the University of Wisconsin-Madison (IRB #2019-1258), University of Chicago Biological Sciences Division (IRB #18-0447), Loyola University Medical Center (IRB #215437), and NorthShore University (IRB #11-0539) Institutional Review Boards with a waiver of informed consent.

### Data Collection

Detailed structured EHR data (patient demographics, vital signs, nursing flowsheets, laboratory results, and billing data) were collected from each health system’s electronic data warehouse. These data were used to retrospectively calculate the eCART score values to identify patients for study inclusion, and the time of first high eCART at or above the 95^th^ percentile for each admission was used to denote the time of clinical deterioration.

In the final study cohort, retrospective manual chart review with expert annotation was performed by trained reviewers at each health system (SR, RI, EM, LN, CN, BDF, JP, and TT). Patient data collected from the chart review was inputted into a Research Electronic Data Capture (REDCap) tool licensed by each institution, and a manual of procedures was created that provided detailed instructions for the chart reviews (19).

The chart review questions were developed by a critical care physician (MMC) and hospitalist (DPE) with experience implementing early warning scores following a review of the literature and discussions with the University of Chicago Medicine’s rapid response team members. The first 20 charts at each site were completed by both chart reviewers as well as the lead investigator at each study site (MMC, CJW, EG, DPE), and a Cohen’s kappa statistic was calculated to establish the level of agreement among the reviewers for the diagnosis causing deterioration. Any disagreements were discussed between reviewers and the site lead as well as with the overall study lead (MC) to increase consistency within and across sites. If the kappa was <0.6, then additional training was provided to the reviewer, and 20 new charts were reviewed by both the site lead and the reviewer. This was repeated until the Cohen’s kappa was ≥ 0.60, and then the reviewer was allowed to proceed with chart reviews independently. Any questions by the reviewer during independent chart review were directed to the site lead for further discussion and to the study lead if uncertainty remained. After 500 charts were completed at each site, another 20 charts were dually reviewed by the reviewers and the site leads, Cohen’s kappa was recalculated, and additional training provided as necessary.

The first question of the chart review determined whether the elevated eCART score was due to a true clinical deterioration episode, which was defined as an underlying medical condition causing true physiologic abnormalities that could lead to critical illness or death if untreated, or a false alarm (e.g., vital sign abnormalities following physical therapy). For patients with a true deterioration episode, the reviewers assessed what the clinicians documented as their initial suspected cause of deterioration at the time of the event as well as what the ultimate cause was determined to be after all the relevant information became available (e.g., diagnostic test results, response to therapy, assessments from specialist consultants, etc.). Up to two primary causes of deterioration could be documented for each deterioration episode. The diagnostic tests and treatments ordered related to the deterioration event were also collected. If the clinicians did not specify any diagnoses as a most likely cause of deterioration, then data were collected on all diagnoses, diagnostic tests, and drug and non-drug interventions performed to treat the clinical deterioration. The final REDCap chart review datasets and structured EHR data from each study site were de-identified and transferred to the University of Wisconsin-Madison for analysis. Free-text diagnoses, tests, and treatments that were documented as comments in the “Other” category by chart reviewers were combined into additional categories as clinically appropriate.

### Statistical Analysis

Descriptive statistics, including the number of false alarms and ultimate causes of deterioration, were calculated from the combined REDCap dataset. Patient characteristics, vital signs, laboratory values, and in-hospital mortality rates were compared across causes of deterioration using Wilcoxon rank sum tests and chi-squared tests, as appropriate for the variable distribution. Causes and outcomes were also compared for conditions present on admission compared to those not present on admission. Descriptive statistics were calculated for diagnostic tests and treatments, and comparisons between patients who survived vs. died during their admission were made. Analysis was performed using Stata Version 16.1 (StataCorp, College Station, TX), and a two-sided p-value of less than 0.05 was considered statistically significant.

## RESULTS

### Study Population

Of 919,319 admissions to the medical-surgical wards during the study period, 91,131 (10%) had at least one eCART value at or above the 95^th^ percentile and were eligible for inclusion. From this cohort, 1,000 randomly selected admissions from each health system (4,000 total) were included in this study (**eFigure 1**). The median age of the study cohort was 69 (IQR 58-82), 49% were female, and 23% were black. A total of 983 (25%) required ICU transfer following their elevated eCART score, and 474 (12%) patients died during their admission. True deterioration events occurred in 2,484 (62%) of the 4,000 included patients based on manual chart review. As shown in **Table 1**, patients with a true deterioration event were older (median age 70 vs. 68 years; p<0.01), more likely to be female (50% vs. 47%; p=0.04), less likely to be black (20% vs. 29%; p<0.01), and had a higher rate of ICU transfer (29% vs. 17%; p<0.01) and in-hospital mortality (14% vs. 8%; p<0.01).

**Table 1:** Comparison of patient characteristics between patients with and without a true deterioration event. Abbreviations: eCART = electronic Cardiac Arrest Risk Triage; LOS = length of stay; ICU = intensive care unit

| **Variable** | **True deterioration events**<br>**(n=2,484)** | **False Alarms**<br>**(n=1,516)** | **p-value** |
| --- | --- | --- | --- |
| Age (years), Median (IQR) | 70 (58, 84) | 68 (57, 79) | <0.01 |
| Female, n (%) | 1,245 (50%) | 709 (47%) | 0.04 |
| Black, n (%) | 492 (20%) | 441 (29%) | <0.01 |
| High eCART score value,<br>Median, IQR | 50 (40, 76) | 49 (39, 78) | 0.64 |
| LOS before high eCART<br>score (hours), Median (IQR) | 20 (6, 67) | 44 (14.5, 111.5) | <0.01 |
| LOS after high eCART<br>score (hours), Median (IQR) | 127 (68, 242) | 103 (44, 215) | <0.01 |
| ICU transfer after high<br>eCART score, n (%) | 721 (29%) | 262 (17%) | <0.01 |
| In-hospital mortality, n (%) | 357 (14%) | 117 (8%) | <0.01 |

ICU = intensive care unit

### Causes of Clinical Deterioration

Sepsis was the most common cause of a true deterioration event (41% of encounters with true deterioration; n=1,021), followed by arrhythmia (19%; n=473), congestive heart failure/volume overload (13%; n=317), hypoxemic respiratory failure (10%; n=256), and electrolyte abnormality (9%; n=231) (**Figure 1**). Most causes of deterioration were present on admission (74%; n=1,816), and the remainder developed during the hospital stay. It was unknown whether the conditions were present on admission for 44 patients, so they were excluded from this calculation. Sepsis was also the most common cause of deterioration that was present on admission (48% of encounters with a true deterioration event present on admission; n=863), followed by arrhythmia (16%; n=286), and congestive heart failure/volume overload (14%; n=256). In contrast, arrhythmia (29% of true deterioration events that were not present on admission; n=183) was the most common cause that developed in the hospital, followed by sepsis (22%; n=140), and hypovolemia (12%; n=72).

**Figure 1:**
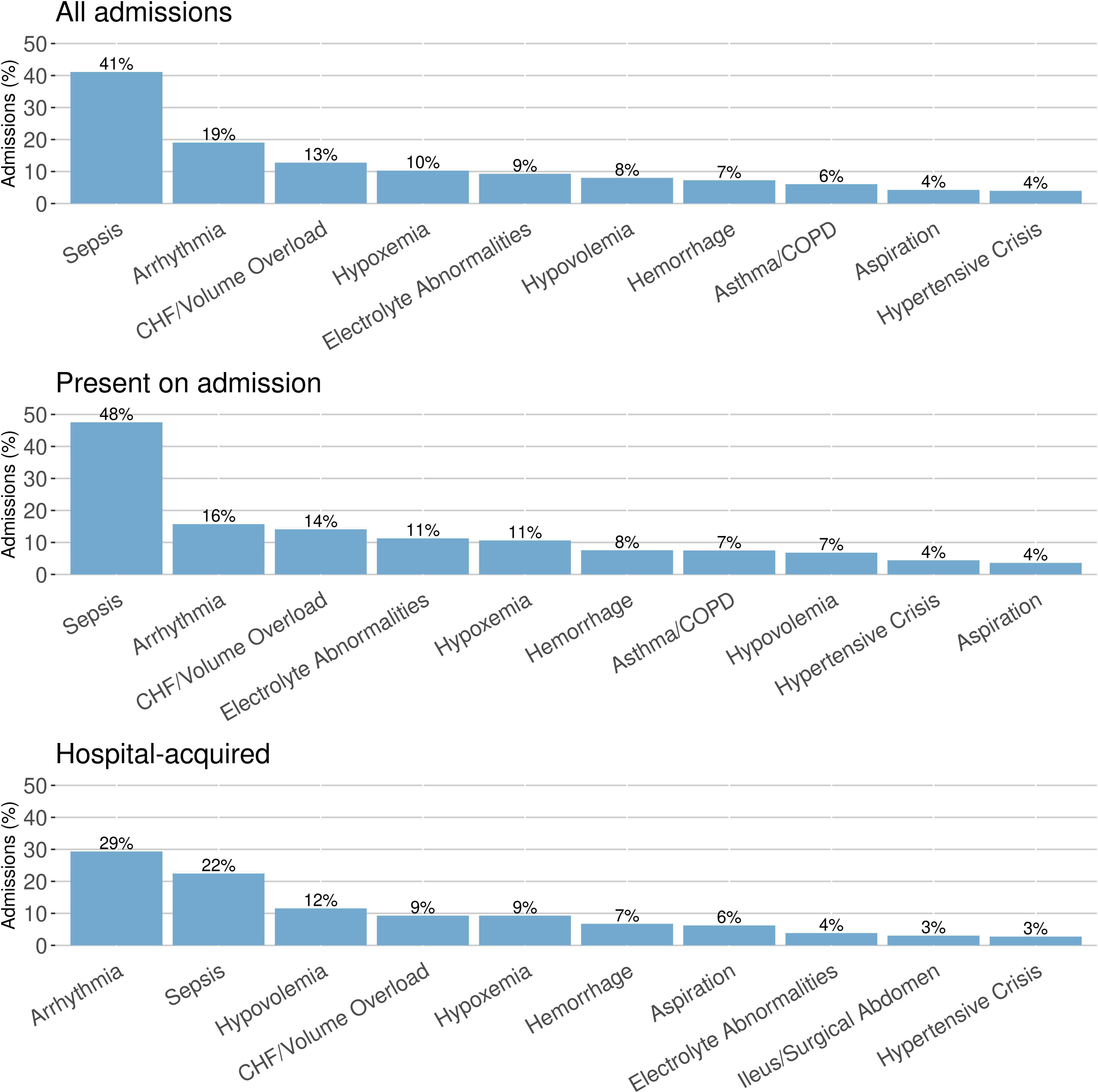
Most common conditions causing true deterioration events overall (top), present on admission (middle), and hospital-acquired (bottom). Abbreviations: CHF = congestive heart failure; COPD = chronic obstructive pulmonary disease

### Outcomes by Underlying Diagnosis

Liver failure had the highest in-hospital mortality rate (41%; n=24), followed by stroke (40%; n=23), hypoxemic respiratory failure (30%; n=78), and pulmonary embolism (30%; n=11) (**Figure 2**). Patients who deteriorated due to conditions that were present on admission had higher in-hospital mortality (15.4%) compared to those deteriorating due to hospital-acquired conditions (10.6%; p=0.003). In contrast, post-sedation hypoventilation (80%; n=4) had the highest rate of ICU transfer, followed by post-operative mechanical complication (64%; n=7), and ileus/surgical abdomen (53%; n=40).

**Figure 2:**
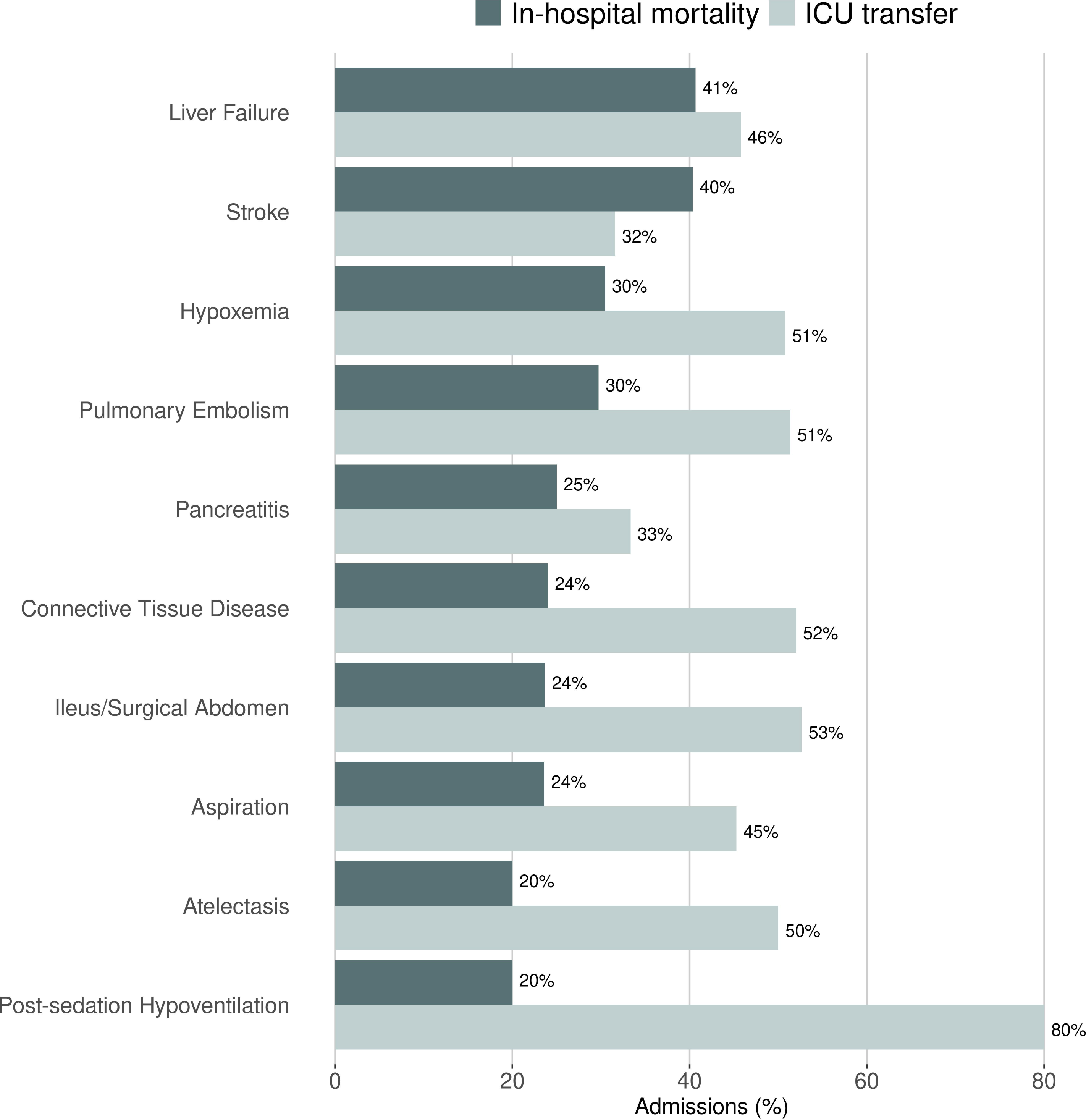
Conditions causing true deterioration events associated with the highest mortality (dark bars) and ICU transfer (light bars) rates.

### Diagnostic Testing and Interventions

Almost all (99%) true deterioration episodes had at least one diagnostic test ordered related to the underlying cause, with a median (IQR) of 3 (2–6) tests ordered. The most common diagnostic tests ordered that helped diagnose the cause of deterioration were complete blood counts (47% of events) followed by chest x-rays (42%), and cultures (40%) (see **Figure 3** for the most common orders and **Supplemental Table 1** for the full list of diagnostic tests). A median (IQR) of 2 (1, 3) non-drug interventions were delivered for each deterioration episode, and the most common non-drug interventions were non-critical care consults (48% of events) followed by critical care consults (23%), and telemetry (21%) (**Figure 3**; **Supplemental Table 2**). A median (IQR) of 2 (1–3) drug interventions were delivered for each deterioration episode, and the most common drug interventions were antimicrobials (46% of events) followed by fluid boluses (34%), and antiarrhythmics (19%) (**Figure 3**; **Supplemental Table 3**).

**Figure 3:**
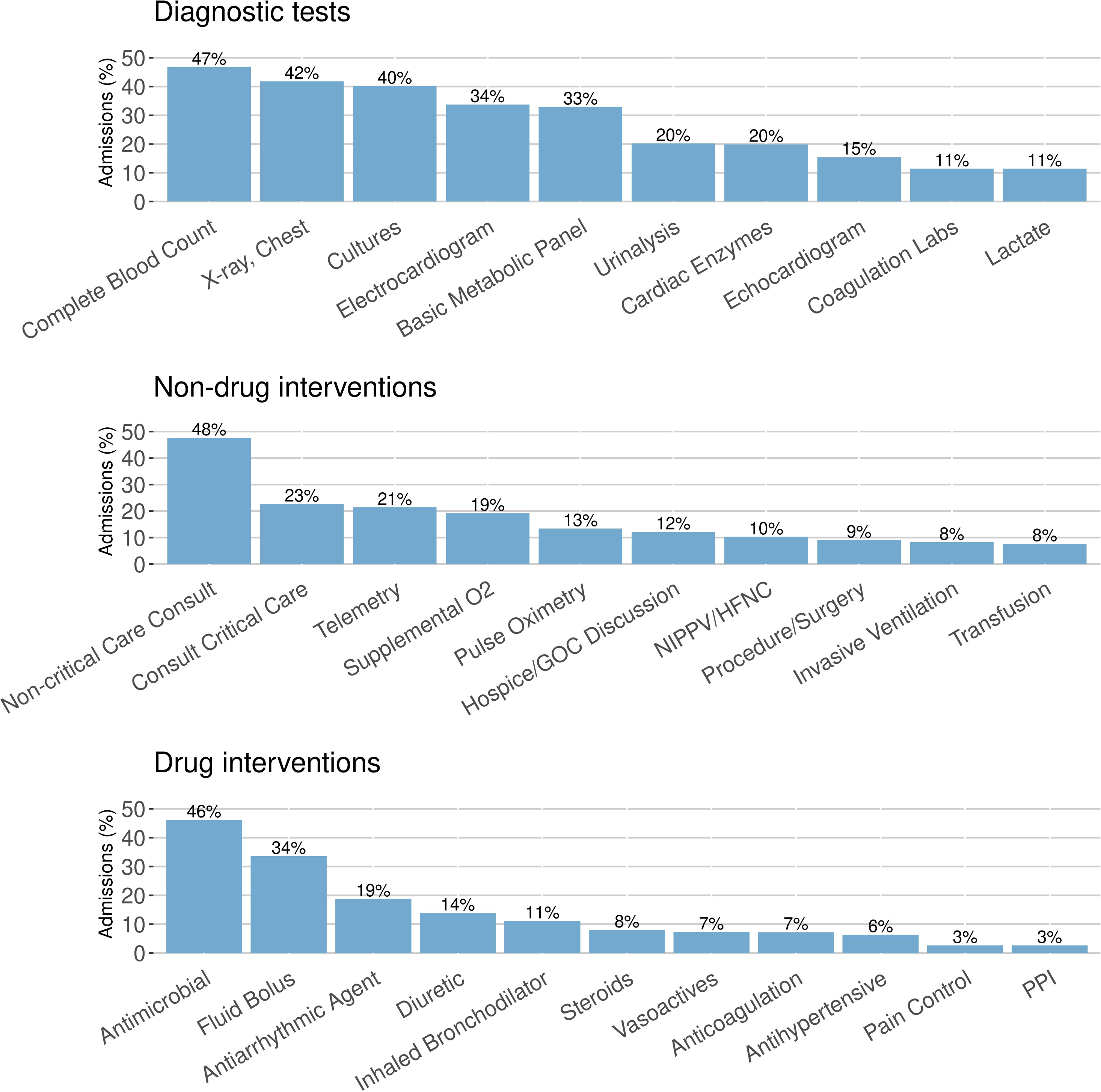
The most common diagnostic tests (top), non-drug interventions (middle), and drug interventions (bottom) ordered related to true deterioration events. Abbreviations: GOC = goals of care; NIPPV = non-invasive positive pressure ventilation; HFNC = high-flow nasal cannula; PPI = proton-pump inhibitor

## DISCUSSION

Using manual chart review of 4,000 patients with clinical deterioration, we determined the most common causes of deterioration, their subsequent outcomes, and the most common diagnostic tests and treatments related to these events. Specifically, we found that sepsis was the most common cause of deterioration, liver failure had the highest mortality, and complete blood counts, antimicrobials, and non-critical care consults were the most common tests and interventions ordered. To our knowledge, this is the first multicenter study that performed detailed chart review to determine the underlying cause of clinical deterioration in high-risk ward patients. These results provide valuable information for clinicians treating these patients and for health systems aiming to create evidence-based treatment pathways for deteriorating patients.

Few studies have investigated causes of clinical deterioration using chart review. For example, in a single-center study Blackwell et al. performed manual chart review on 457 patients transferred to the ICU from an adult acute care cardiac and cardiovascular surgery ward (16). They found that respiratory instability alone and respiratory instability plus suspected sepsis were the most common causes of ICU transfer. This study differs from ours in that it was in one cardiac medical-surgical ward and the results focused on patients admitted to the ICU as opposed to all high-risk patients with clinical deterioration on the wards as identified by an early warning score. Lyons and colleagues published results from the first 402,023 rapid response team calls in the Get with the Guidelines-Medical Emergency Team registry, which used staff data entry into an online reporting form (15). They found that respiratory and cardiac triggers were the most common reasons for team activation and that additional monitoring and fluids were the most common orders. Another smaller study of 1,151 rapid response team calls by White et al. found that hypotension, decreased level of consciousness, and oxygen desaturation were the most common call triggers (18). These prior studies focused specifically on rapid response team calls and physiological reasons for activations, which is different from the focus of our current study on all high-risk patients identified by a validated early warning score. The lack of other large-scale studies on the causes and treatments for deteriorating ward patients likely relates to the time-consuming nature of manual chart reviews, which are important to determine which diagnoses and treatments are the cause of the event.

The fact that sepsis was the most common cause of deterioration is consistent with prior work by Liu and colleagues suggesting that infections played a key role in up to 50% of deaths in the hospital (20). Further our group previously found that 27.5% of hospitalized patients experienced at least one episode of infection during their admission (21). In this current study, sepsis was the most common cause of deterioration, while liver failure was the deadliest cause, with a mortality rate of 41%. We also found that the causes of deterioration were different based on whether they were present on admission. Sepsis was the common condition present on admission, while arrythmias were the most common condition that was newly acquired during the hospitalization. In-hospital mortality also varied based on whether or not the condition was present on admission. These findings provide valuable diagnostic and prognostic information for clinicians at the bedside responding to a newly deteriorating patient.

Diagnostic testing was ordered to determine the cause of deterioration in almost all cases. Complete blood counts, chest x-rays, cultures, and electrocardiograms were the most common orders, which correspond to some of the most common causes of deterioration, as expected. Interventions also corresponded to these causes, with antimicrobials, fluid boluses, and antiarrhythmics being the most ordered medications. Non-critical care consultants were also requested in half of true deterioration events. These results can assist hospitals with resource planning for these events, as institutions are increasingly developing care pathways to standardize and streamline patient care. Further, this information could also be used when developing case scenarios for clinician training in response to clinical deterioration.

Our study has several strengths. Most importantly, we determined the causes, diagnostic tests, and interventions related to clinical deterioration using manual chart review by clinicians as opposed to using billing codes. These clinicians had access to the entire medical record from the admission to make these determinations, and agreement between reviewers was high. Moreover, there was an objective time zero because of the combination of an early warning score with manual chart review, which allowed for determining the deterioration cause related to the score elevation. In addition, our study was performed at four different health systems, which included academic, suburban teaching, and community hospitals, which should increase generalizability. Finally, to our knowledge, this is the first study of its kind performed in a general ward population, which can provide insights to improve the care of hospitalized patients.

In addition to these strengths, our study also has limitations. First, although the chart reviewers had access to the results of diagnostic testing and other information from the chart, diagnostic uncertainty is still possible, which could affect the results. However, agreement between reviewers was high, suggesting that certainty was possible in many cases. This is consistent with our prior chart review study focused on infection that explicitly measured reviewer uncertainty and found that the uncertain category (“possible infection”) was by far the least commonly selected. Second, our study was performed at four health systems in the Midwest United States, so the results may not be generalizable to other geographical regions. Finally, our cohort was identified using one specific early warning score, and results may differ if other scores are used. For example, eCART has been shown to have increased sensitivity and specificity compared to the commonly used MEWS, which would have resulted in more patients being excluded from the main analyses due to being false positives if MEWS were used (5).

## CONCLUSIONS

In conclusion, we found that sepsis was the most common cause of clinical deterioration, while liver failure had the highest mortality. Complete blood counts and chest x-rays were the most common diagnostic tests ordered, and antimicrobials and fluid boluses were the most common medications ordered to treat the cause of deterioration. These results provide important insights for clinical decision-making at the bedside, training of rapid response teams, and the development of institutional treatment pathways for clinical deterioration.

## Data Availability

The data utilized in this article cannot be shared publicly because of legal and regulatory restrictions. These data were obtained from four hospital systems after our research protocol was reviewed by IRBs from each hospital, and our data use agreements do not permit sharing due to the granular nature of the data.

## ACKNOWLEDGEMENTS

None

## Author contributions

MMC takes full responsibility for the content of the manuscript. MMC and DPE conceptualized the study. KAC, RI, and TQ conducted statistical analysis of the data. MMC wrote the first draft of the manuscript and revised subsequent versions. All authors contributed to the interpretation of data, reviewed and edited the initial drafts, and approved the final manuscript.

