## Supplementary material for "Causes, Diagnostic Testing, and Treatments Related to Clinical Deterioration Events among High-Risk Ward Patients": ONLINE DATA SUPPLEMENT

**eFigure 1:** Study patient flow diagram.

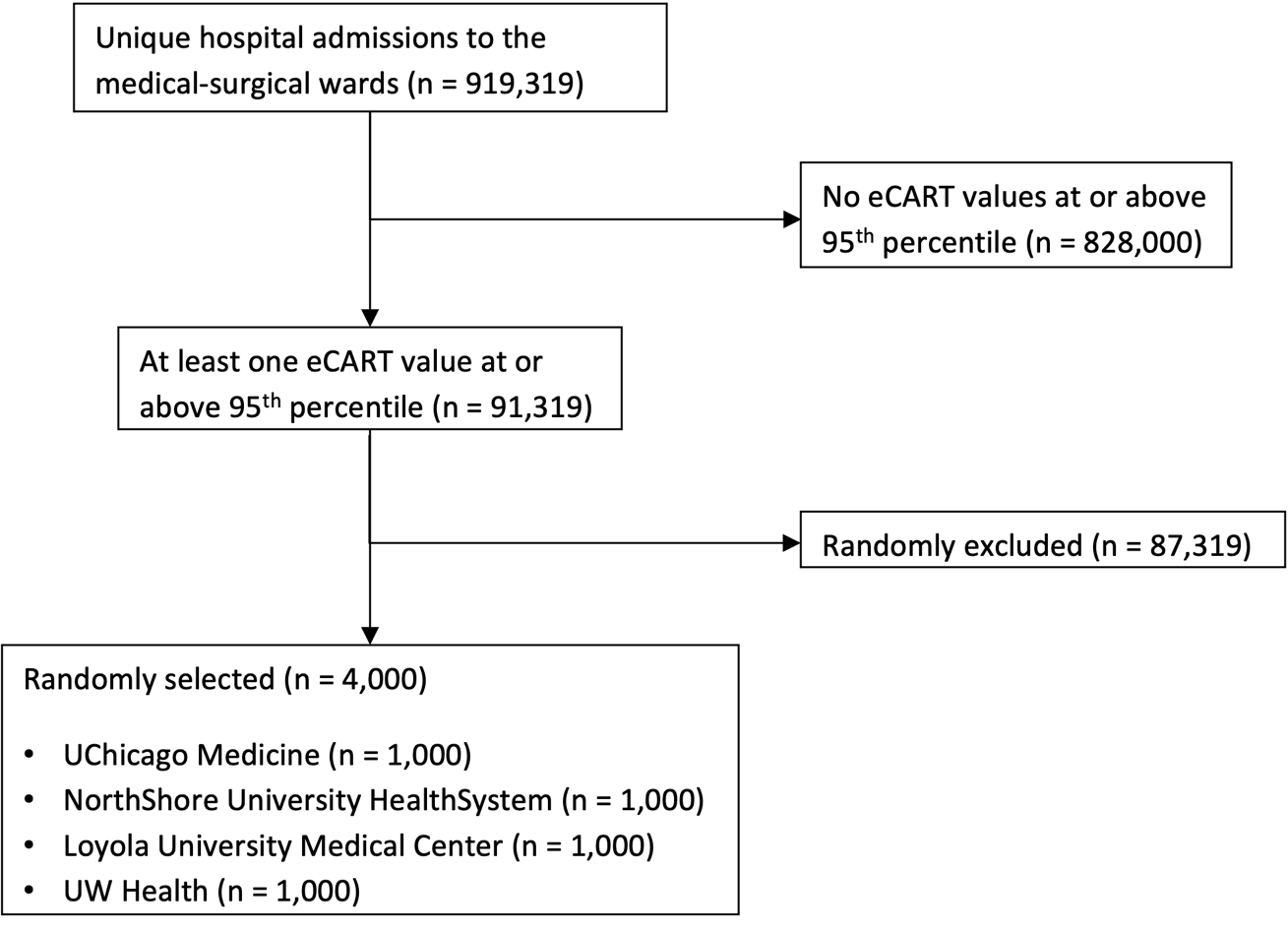

**eTable 1:** Full list of diagnostic tests ordered and their frequency for patients with true deterioration events (n=2,484).

| **Diagnostic test** | **Encounters, n (%)** |
| --- | --- |
| Complete Blood Count | 1,161 (47%) |
| X-ray, Chest | 1,038 (42%) |
| Cultures | 998 (40%) |
| Electrocardiogram | 838 (34%) |
| Basic Metabolic Panel | 818 (33%) |
| Urinalysis | 502 (20%) |
| Cardiac Enzymes | 494 (20%) |
| Echocardiogram | 384 (15%) |
| Coagulation Labs | 284 (11%) |
| Lactate | 284 (11%) |
| CT, Abdomen/Pelvis | 281 (11%) |
| CT, Chest | 252 (10%) |
| Arterial Blood Gas | 223 (9%) |
| Liver Function Tests | 188 (8%) |
| CT, Head | 155 (6%) |
| Abdominal Ultrasound | 82 (3%) |
| X-ray, Abdomen/Pelvis | 81 (3%) |
| Endoscopy | 73 (3%) |
| Respiratory Viral Panel | 67 (3%) |
| Extremity Dopplers | 66 (3%) |
| Glucose | 62 (2%) |
| CRP | 46 (2%) |
| ESR | 45 (2%) |
| Electroencephalogram | 44 (2%) |
| Paracentesis | 39 (2%) |
| Thoracentesis | 37 (1%) |
| X-ray, Other | 32 (1%) |
| Bronchoscopy | 30 (1%) |
| CT, Other | 24 (1%) |
| Thyroid Studies | 24 (1%) |
| MRI, Head | 23 (1%) |
| FOBT | 20 (1%) |
| MRI, Other | 19 (1%) |
| Lipase/Amylase | 18 (1%) |
| Ammonia | 17 (1%) |
| Cardiac Cath | 16 (1%) |
| LP | 13 (1%) |
| Hemolysis Labs | 11 (0.4%) |
| VQ Scan | 11 (0.4%) |
| Procal | 10 (0.4%) |
| Creatinine Kinase | 8 (0.3%) |
| PFTs | 8 (0.3%) |
| Swallow Eval | 8 (0.3%) |
| ANA Panel | 6 (0.2%) |
| B12/Folate | 5 (0.2%) |
| C3, C4 Complement | 5 (0.2%) |
| Cortisol | 5 (0.2%) |
| HIV/HCV/Hepatitis Labs | 5 (0.2%) |
| Iron Studies | 5 (0.2%) |
| Cardiac MRI | 4 (0.2%) |
| D-dimer | 4 (0.2%) |
| Stress Test | 4 (0.2%) |
| UDS | 4 (0.2%) |
| Ultrasound, Other | 4 (0.2%) |
| Carotid Imaging | 2 (0.1%) |
| Ketones | 2 (0.1%) |
| MRCP | 2 (0.1%) |
| BM Biopsy | 1 (0.04%) |
| Lipid Panel | 1 (0.04%) |
| MRI, Abdomen | 1 (0.04%) |

**eTable 2:** Full list of non-drug interventions ordered and their frequency for patients with true deterioration events (n=2,484).

| **Non-drug intervention** | **Encounters, n (%)** |
| --- | --- |
| Non-critical Care Consult | 1,183 (48%) |
| Consult Critical Care | 562 (23%) |
| Telemetry | 532 (21%) |
| Supplemental O2 | 476 (19%) |
| Pulse Oximetry | 333 (13%) |
| Hospice/GOC Discussion | 302 (12%) |
| NIPPV/HFNC | 255 (10%) |
| Procedure/Surgery | 225 (9%) |
| Invasive Ventilation | 204 (8%) |
| Transfusion | 191 (8%) |
| Central Line | 171 (7%) |
| Continuous Monitoring, Other | 140 (6%) |
| Suctioning | 62 (2%) |
| Dialysis | 59 (2%) |
| Thoracentesis | 56 (2%) |
| Chest Physiotherapy | 47 (2%) |
| Cardioversion | 44 (2%) |
| Paracentesis | 27 (1%) |
| ICD/Pacemaker | 13 (1%) |
| Ablation | 11 (0.4%) |
| NG Tube | 8 (0.3%) |
| New Foley | 7 (0.3%) |
| Arterial Line | 6 (0.2%) |
| Bag-valve Mask | 6 (0.2%) |
| Bronchoscopy | 6 (0.2%) |
| DHT | 4 (0.2%) |
| EGD | 3 (0.1%) |
| Incentive Spirometry | 3 (0.1%) |
| Remove Line | 3 (0.1%) |
| Enemas | 2 (0.1%) |
| Monitoring Apnea | 2 (0.1%) |

**eTable 3:** Full list of drug interventions ordered and their frequency for patients with true deterioration events (n=2,484).

| **Drug intervention** | **Encounters, n (%)** |
| --- | --- |
| Antimicrobial | 1,147 (46%) |
| Fluid Bolus | 834 (34%) |
| Antiarrhythmic Agent | 466 (19%) |
| Diuretic | 346 (14%) |
| Inhaled Bronchodilator | 278 (11%) |
| Steroids | 200 (8%) |
| Vasoactives | 182 (7%) |
| Anticoagulation | 180 (7%) |
| Antihypertensive | 159 (6%) |
| Pain Control | 66 (3%) |
| PPI | 66 (3%) |
| Aspirin | 63 (3%) |
| Insulin | 48 (2%) |
| Dextrose | 45 (2%) |
| Sedative | 40 (2%) |
| Lactulose | 23 (1%) |
| Anti-epileptic | 22 (1%) |
| Antihistamine | 21 (1%) |
| Kayexalate | 21 (1%) |
| Narcan | 21 (1%) |
| Electrolytes | 20 (1%) |
| Nitroglycerin | 15 (1%) |
| Albumin | 13 (1%) |
| Epinephrine | 12 (0.5%) |
| Bicarb | 9 (0.4%) |
| Calcium | 9 (0.4%) |
| Octreotide | 9 (0.4%) |
| Benzos | 8 (0.3%) |
| Acetylcysteine | 7 (0.3%) |
| Immunosuppressives | 6 (0.2%) |
| Beta Blocker | 5 (0.2%) |
| Chemo | 5 (0.2%) |
| D/C Antihypertensives | 5 (0.2%) |
| G-CSF | 5 (0.2%) |
| Rifaximin | 5 (0.2%) |
| Vitamin K | 5 (0.2%) |
| Midodrine | 4 (0.2%) |
| Salt Tabs | 3 (0.1%) |
| Atropine | 2 (0.1%) |
